# A Machine Learning Based Causal Interface for Time Varying Environmental Predictors of Substance Use Initiation in the ABCD Study

**DOI:** 10.64898/2026.04.15.26350988

**Authors:** Mengman Wei, Lasya Yadlapati, Qian Peng

## Abstract

**Background:** The Adolescent Brain Cognitive Development (ABCD) Study^®^ offers rich longitudinal data on environmental, genetic, and other factors related to substance use initiation. Classical marginal structural models (MSMs) require choosing which covariates to include in propensity models, but this choice is difficult in the presence of hundreds of correlated predictors.

**Methods:** We analyzed longitudinal data from the Adolescent Brain Cognitive Development Study using a European-ancestry unrelated cohort, in which each individual contributed repeated observations over time. Interval-level binary outcomes were defined for initiation of alcohol, nicotine, cannabis, and any substance, restricting analyses to participants at risk prior to initiation. All predictors were constructed as lagged variables to preserve temporal ordering. We implemented a two-stage machine learning–based causal framework. First, we performed graph discovery using a “Granger-inspired lagged predictive modeling” approach, applying elastic-net logistic regression to identify predictive relationships between lagged environmental variables and future initiation outcomes. Robust candidate edges were selected using subject-level bootstrap stability selection. Second, we estimated adjusted effect sizes for stable edges using double machine learning (DML)-style partialling-out with cross-fitting. For each candidate predictor, we defined the treatment as the lagged variable of interest and adjusted for high-dimensional lagged covariates. Cross-fitting with group-based splitting accounted for within-subject dependence, and nuisance functions were estimated using random forest models. Cluster-robust standard errors were used for inference.

**Results:** Across analyses, behavioral and environmental predictors showed stronger and more consistent associations with substance-use initiation than polygenic risk scores (PRSs). In the graph-discovery analysis, stable edges were mainly related to rule-breaking behavior, sensation seeking, resiliency, sleep, screen-related measures, and family- or parent-related variables. PRS variables were less consistently selected and did not appear as dominant predictors. Cannabis initiation showed strong stable links with parent-reported rule-breaking behavior and behavioral symptoms. Nicotine initiation was linked to sensation seeking, behavioral symptoms, and screen-related measures. DML-style effect estimates were modest in magnitude but supported positive associations for several stable predictors, including rule-breaking behavior for cannabis initiation and sensation seeking, Brief Problem Monitor (BPM) behavioral items, and screen-related measures for nicotine initiation.

**Conclusions:** In this European-ancestry unrelated cohort, adolescent substance-use initiation was more consistently associated with modifiable behavioral and environmental factors than with PRS variables. The findings suggest that cannabis and nicotine initiation share some risk factors but also have distinct predictor profiles. These results highlight the importance of longitudinal, multi-method approaches for identifying early risk patterns and suggest that prevention strategies may benefit from focusing on behavioral regulation, sensation seeking, family context, sleep, and screen-related behaviors.

## Introduction

Substance use often begins during adolescence, a developmental period when curiosity, risk-taking, and social influences increase while the brain is still maturing. Initiation of alcohol, nicotine, or cannabis use at earlier ages is consistently associated with a higher risk of later substance use disorders and other adverse outcomes, making prevention and early identification of risk a major public health priority [1].

Risk for initiation is multifactorial and spans multiple domains, including individual behavior and mental health, family environment and parenting, peer context, school experiences, and neighborhood and socioeconomic conditions. These influences can change over time, and their timing may matter. For example, changes in peer exposure or parental monitoring during a specific developmental window may be more predictive of near-term initiation than baseline-only measures. Large longitudinal cohorts therefore provide an opportunity to study both whether initiation occurs and when it occurs, using modeling frameworks that respect time ordering [2, 3]. The Adolescent Brain Cognitive Development (ABCD) Study^®^ is particularly well suited for this work because it is a large, diverse, multi-site longitudinal cohort that follows youth from late childhood into adolescence with rich repeated assessments across psychosocial,environmental, and health domains [4, 5].

Methodologically, two complementary goals are important. First, we need a principled way to screen thousands of time-varying predictors to identify candidate directional relationships that respect temporal precedence (past → future). A common approach is Granger-style analysis, where a predictor is considered informative if its past values improve the prediction of a later outcome, with directionality arising from time ordering rather than simultaneity [6, 7]. In high-dimensional settings, sparse models such as elastic-net logistic regression can perform variable selection while handling correlated predictors, and stability selection (repeated resampling or subsampling) can help distinguish robust signals from chance selections [8, 9, 10, 11, 12]. Second, after identifying robust candidate edges, we want effect-size estimates that adjust for confounding from other past variables. Double/debiased machine learning (DML) provides a framework for estimating low-dimensional effects while flexibly modeling high-dimensional nuisance relationships with machine learning and using cross-fitting to reduce overfitting bias [13, 14, 15, 16].

In this work, we combine these ideas into a two-stage pipeline:

1. graph discovery using lagged predictors, sparse logistic regression, and subject-level bootstrap stability to propose directional edges; and
2. effect estimation for stable edges using DML-style partialling-out with cross-fitting, with machine-learning nuisance models and cluster-robust uncertainty to account for repeated measures within individuals (Figure 1). Together, this approach aims to provide an interpretable causal interface that highlights which time-ordered factors are most consistently linked to subsequent initiation risk, and estimates their adjusted associations under standard causal assumptions.

**Figure 1.**
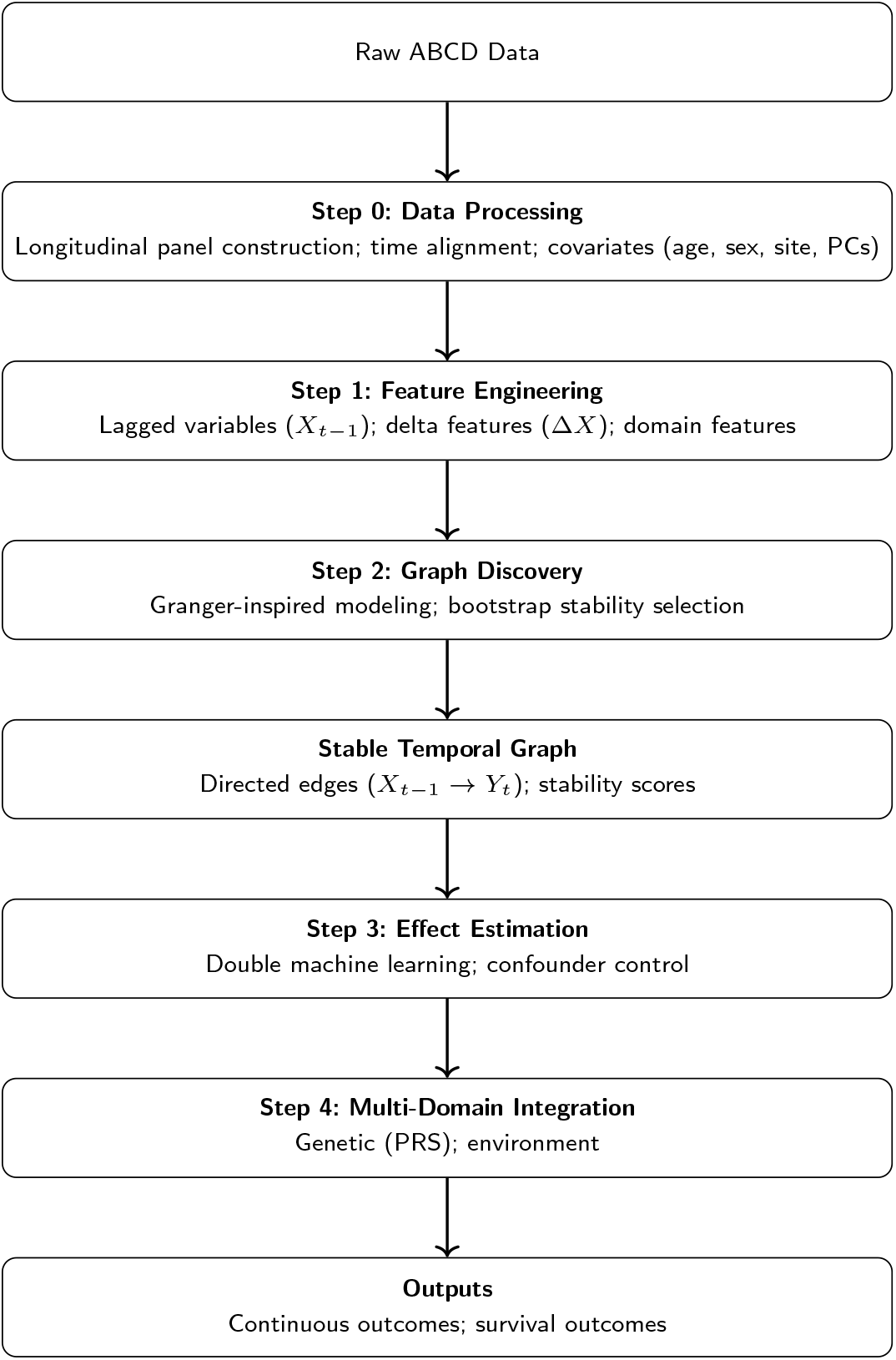
Overview of the multi-stage modeling pipeline. This study uses a structured pipeline to integrate longitudinal, genetic, and environmental data from the ABCD Study. First, raw data are processed into a time-aligned longitudinal panel with covariate adjustment. Next, feature engineering constructs lagged and change (delta) variables to capture temporal dynamics. A graph discovery step then identifies stable temporal relationships using Granger-inspired lagged predictive modeling with bootstrap stability selection. Based on the resulting temporal graph, causal effects are estimated using double machine learning while controlling for confounders. Finally, features from multiple domains, including polygenic risk scores, and environmental exposures are integrated to model both continuous behavioral outcomes and time-to-event outcomes related to substance use initiation.

## Methods

### Cohort

We used data from the Adolescent Brain Cognitive Development (ABCD) Study^®^ release 5.1 [5]. Participants were eligible for inclusion if they had genome-wide genotyping data that passed quality control, at least one visit with substance-use data, and complete core covariate information, including sex, age, study site, and ancestry principal components.

From the full ABCD baseline cohort of 11,869 participants, we restricted all analyses to unrelated participants of European genetic ancestry. Participants were also required to have available polygenic risk scores, covariates, and longitudinal substance-use initiation data. This restriction was applied to reduce potential confounding from population stratification and genetic relatedness. It also ensured that the analytic sample was consistent with the European-ancestry framework used to construct the polygenic risk scores.

The final European-ancestry unrelated analytic cohort included 2,366 participants. At baseline, the mean age was 9.49 years, with a standard deviation of 0.51 years. The cohort included 1,247 male participants and 1,119 female participants, corresponding to 52.7% male and 47.3% female.

We examined four time-to-event outcomes: alcohol initiation, nicotine initiation, cannabis initiation, and any substance initiation. During follow-up, 964 participants initiated alcohol use, 151 initiated nicotine use, 100 initiated cannabis use, and 1,027 initiated any substance use. These corresponded to event rates of 40.7% for alcohol initiation, 6.4% for nicotine initiation, 4.2% for cannabis initiation, and 43.4% for any substance initiation.

Sex-specific event rates were generally similar for alcohol, cannabis, and any substance initiation. Alcohol initiation occurred in 41.6% of female participants and 39.9% of male participants. Cannabis initiation occurred in 3.9% of female participants and 4.5% of male participants. Any substance initiation occurred in 44.1% of female participants and 42.8% of male participants. Nicotine initiation was slightly higher among female participants, occurring in 7.6% of female participants compared with 5.3% of male participants. Additional details are provided in our previous work [17].

### Study design and data format (longitudinal panel data)

We analyzed longitudinal data in which each participant contributes multiple observations over time, following a standard panel data framework [18, 19]. Each row in the dataset corresponds to one discrete follow-up interval (indexed by step).

For each substance outcome (e.g., alcohol, nicotine, cannabis), we defined an interval-level binary outcome indicating whether initiation occurred during that interval (1 = initiated in that interval, 0 = did not initiate in that interval). Because participants are only at risk of initiation prior to first use, we additionally defined an at-risk (validity) indicator *m*_*_ for each outcome. Specifically, *m*_*_ = 1 indicates that the participant is still at risk and the corresponding row is included for that outcome, whereas *m*_*_ = 0 indicates that the row is excluded for that outcome.

To ensure proper temporal ordering and avoid information leakage, all candidate predictors were constructed using lagged values from prior intervals [18]. Lagged variables were denoted using the suffix `L`*k*, where *k* represents the lag length (e.g., `L1` corresponds to the previous interval and `L2` to two intervals earlier). The analysis focused exclusively on these lagged predictors so that all predictors temporally precede the outcomes.

### Step 1: Graph Discovery Using Lagged Prediction and Bootstrap Stability

We first identified candidate directional relationships from past predictors to future outcomes using a Granger-inspired lagged predictive modeling approach, in which a predictor is considered potentially influential if its past values improve the prediction of a future outcome [7].

#### Eligibility criteria

For each outcome, we included only rows that satisfied two conditions: (1) the participant was at risk for that outcome, and (2) at least one prior interval was available. To ensure stable estimation, we further excluded targets with fewer than 500 eligible rows, extremely low or extremely high prevalence, or no variation in the outcome.

#### Candidate predictors

The predictor set consisted of all lagged variables with suffixes `L1` through `Lmax lag`. Identifier and bookkeeping fields were excluded. Additional details are provided in our previous work [17].

#### Preprocessing

All predictors were converted to numeric format. Infinite values were treated as missing. Missing values were imputed using the column median, with a fallback value of 0 when necessary. Predictors were then standardized (z-scored) to ensure comparability across variables.

#### Sparse predictive model

For each outcome, we fit an elastic-net logistic regression model to predict the interval-level outcome from lagged predictors. Elastic-net regularization promotes sparsity and handles correlated predictors effectively [12]. Class weighting was applied to account for class imbalance.

#### Bootstrap stability selection

To assess robustness, we repeated model fitting across bootstrap samples drawn at the subject level. For each bootstrap replicate, we recorded predictors with non-zero coefficients. For each predictor– outcome pair, we computed a stability score defined as the proportion of bootstrap samples in which the predictor was selected. This procedure follows the principles of stability selection for robust variable identification [11].

#### Outputs of discovery

This step produced a set of candidate directed edges (predictor → outcome) with associated bootstrap stability scores, along with ranked coefficient summaries from the fitted models.

### Step 2: Effect estimation for stable edges using DML-style partialling-out with cross-fitting

After identifying candidate edges, we estimated adjusted effect sizes using DML-style partialling-out with cross-fitting, a framework for adjusted (and causal under assumptions) estimation with machine learning–based nuisance models [20].

#### Stable edge selection

We retained edges exceeding a predefined stability threshold (e.g.,≥ 0.6) and limited the number of edges per outcome.

#### Estimation dataset

Effect estimation was conducted using training and validation splits, excluding the test set. For each outcome, only rows with *m*_*_ = 1 and step ≥ 1 were included.

#### Treatment, outcome, and covariates

- Outcome: interval-level outcome (*y*_*_)
- Treatment: lagged predictor
- Covariates: all lagged variables up to the same lag order (excluding the treatment)

All covariates were numerically encoded, imputed, and standardized.

#### Cross-fitting

To account for repeated measures within individuals, we used group-based cross-validation (GroupKFold) splitting by participant ID. Cross-fitting reduces overfitting bias in causal estimation [20].

#### Nuisance models

We estimated:

- Outcome model: 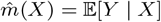
- Treatment model: *ĝ*(*X*) = E[*D* | *X*]

using random forest regression [21].

Residuals were computed as:

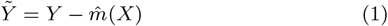

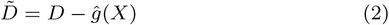

#### Effect estimation

The adjusted effect was estimated by regressing the residualized outcome on the residualized treatment:

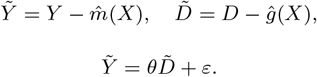

The coefficient *θ* represents the adjusted effect on the probability scale.

### Uncertainty estimation

Because observations are clustered within individuals, we computed cluster-robust standard errors using a sandwich estimator [22].

### Output

This step produced effect estimates, standard errors, and confidence intervals for each retained edge.

### Graph Discovery Summary

The graph discovery step identifies temporally ordered predictive relationships using lagged variables and bootstrap stability. While this step suggests directional associations, it does not establish causality. The DML step provides adjusted effect estimates under standard causal assumptions, including correct temporal ordering and adequate control of confounding through lagged covariates. Together, these steps could yield a robust and interpretable framework for identifying and quantifying candidate risk pathways.

## Results

### Graph Discovery and Stability Selection

We first used longitudinal graph discovery to identify candidate time-varying relationships between baseline and follow-up predictors and later substance-use initiation. Because a single learned graph can be unstable, we used a bootstrap-based stability-selection approach. In each bootstrap run, the graph-learning procedure was repeated, and each directed edge was recorded. We then calculated a stability score for each edge, defined as the proportion of successful bootstrap runs in which that edge was selected.

After graph discovery, we removed technical and adjustment variables from the reported results, including age, sex, site/scanner variables, genetic principal components, time variables, and outcome-label variables. This filtering step allowed the final figures and tables to focus on interpretable predictors, such as behavioral, family, environmental, and genetic-risk variables.

### Overview of Stability Patterns

The stability-selection results showed that several predictors were repeatedly selected across bootstrap samples, suggesting that the discovered graph contained reproducible longitudinal structure rather than only unstable associations. The strongest and most consistent signals were mainly from behavioral and environmental variables.

Across outcomes, several predictors appeared in more than one target-specific graph (Figure 2). The most shared predictors included parent-reported rule-breaking behavior, youth resiliency, family conflict or monitoring-related measures, sleep-related variables, and life-event affect scores. Parent-reported rule-breaking behavior showed the highest maximum stability, with stability reaching 0.915 for cannabis initiation. Youth resiliency also showed high stability, reaching 0.890 for cannabis initiation. Other predictors, including sleep duration and family or environmental measures, showed moderate stability across more than one outcome.

**Figure 2.**
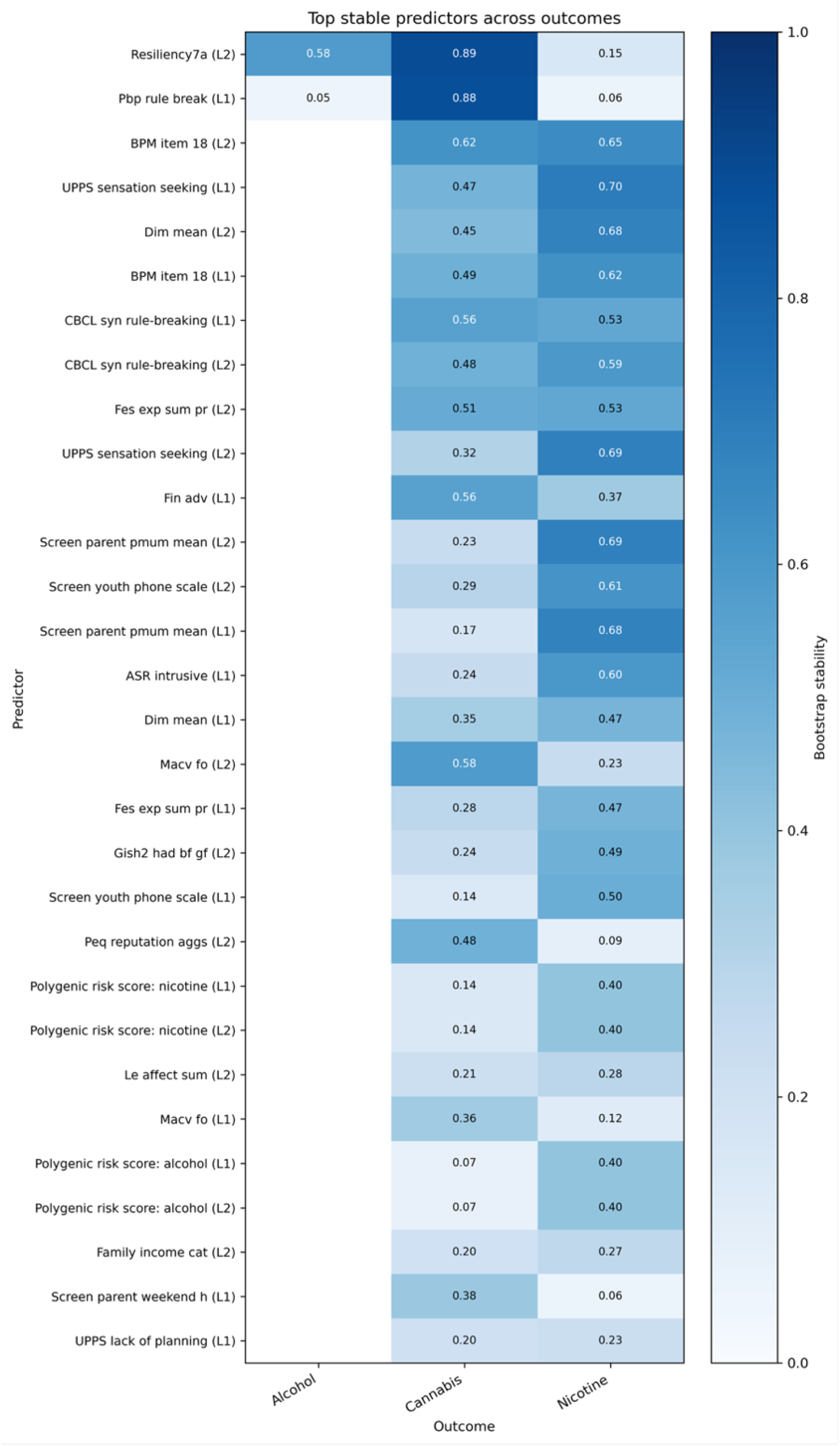
Heatmap of top stable predictors across outcomes. Each cell shows the bootstrap stability score for a predictor–outcome pair. Higher values indicate that the edge was selected more consistently across bootstrap runs. Predictors that appear across multiple outcomes may represent shared risk factors.

In contrast, polygenic risk scores showed weaker and less consistent stability. Alcohol, nicotine, and any-substance polygenic risk scores appeared in some stable-edge summaries, but their maximum stability values were lower than those of the leading behavioral predictors. Cannabis polygenic risk scores showed very low stability in the displayed results. This pattern is consistent with the earlier Cox and marginal structural model results [17], in which behavioral and environmental predictors were more consistently important than polygenic risk score variables.

Together, these results suggest that adolescent substance-use initiation was more strongly connected to behavioral and environmental risk patterns than to genetic-risk scores alone.

### Top Predictors by Outcome

When the stable edges were examined by outcome, the patterns differed across substances, suggesting that each substance-use outcome had a partly distinct risk profile (Figure 3).

**Figure 3.**
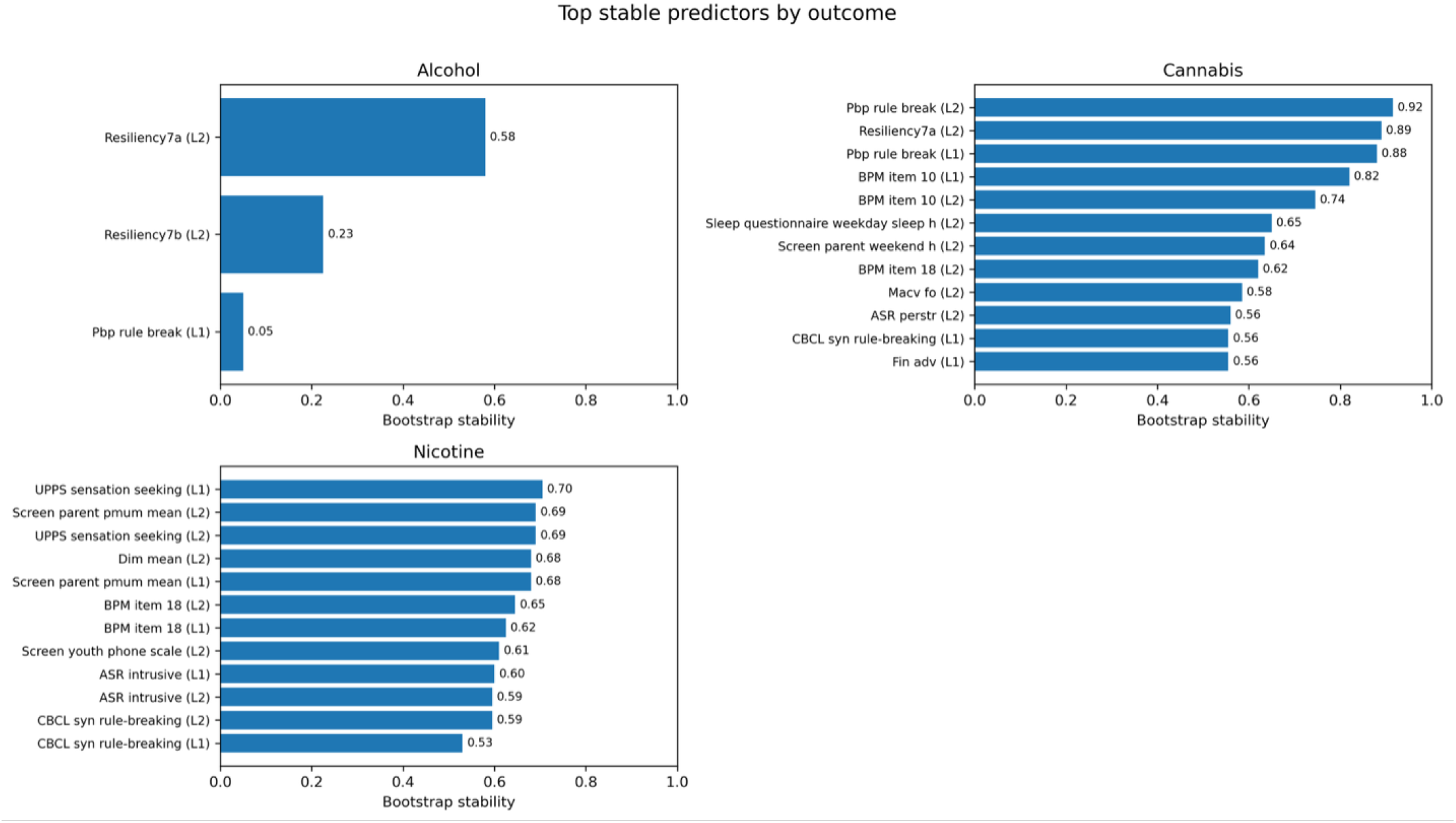
Top stable predictors by outcome. For each substance-use outcome, predictors are ranked by bootstrap stability. Cannabis and nicotine initiation had several highly stable predictors, whereas alcohol initiation showed a smaller set of stable predictors.

**Figure 4.**
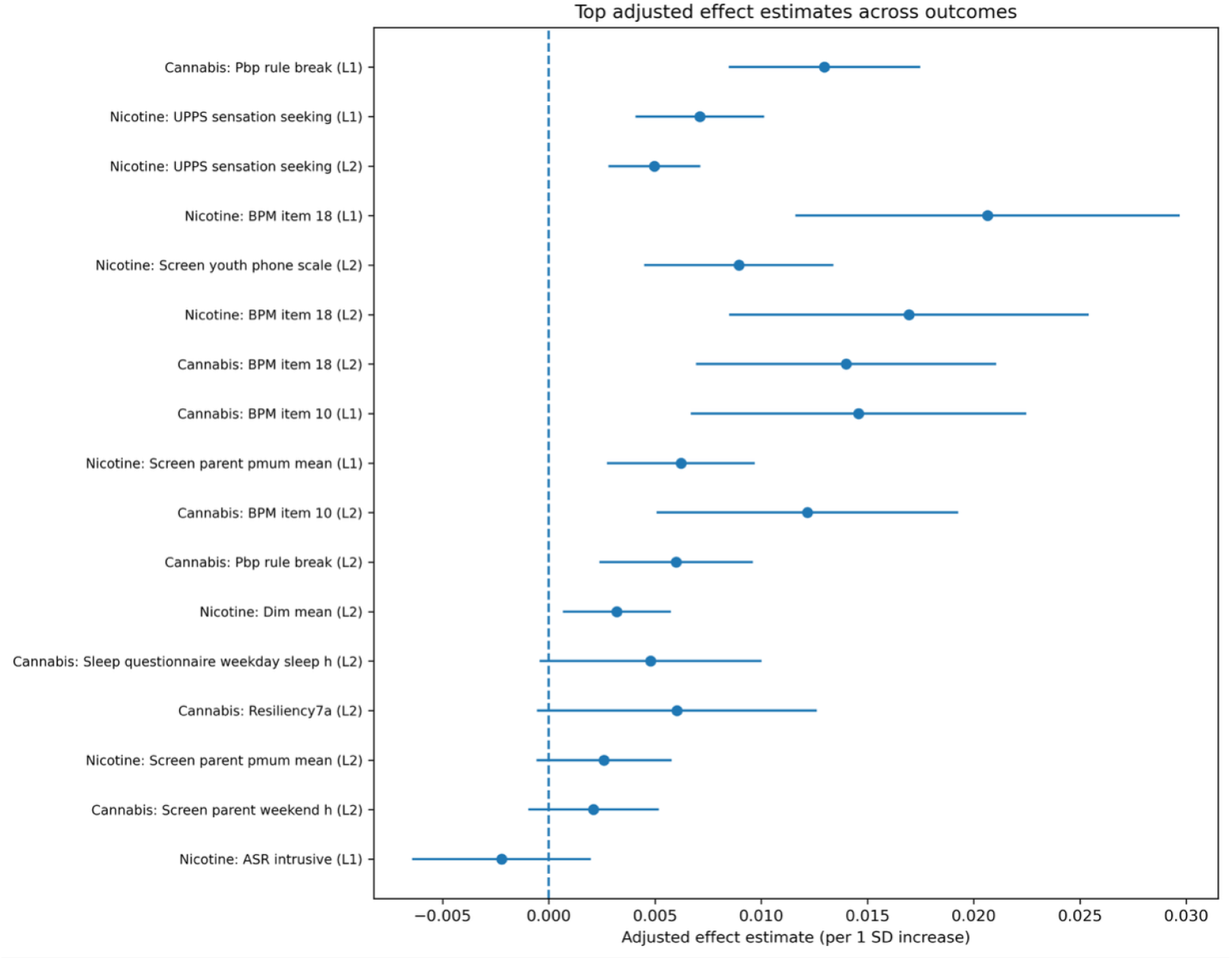
Top DML-style effect estimates across outcomes. Points show the estimated effects per 1-standard-deviation increase in the predictor. Horizontal lines show 95% confidence intervals. The dashed vertical line indicates zero effect.

**Figure 5.**
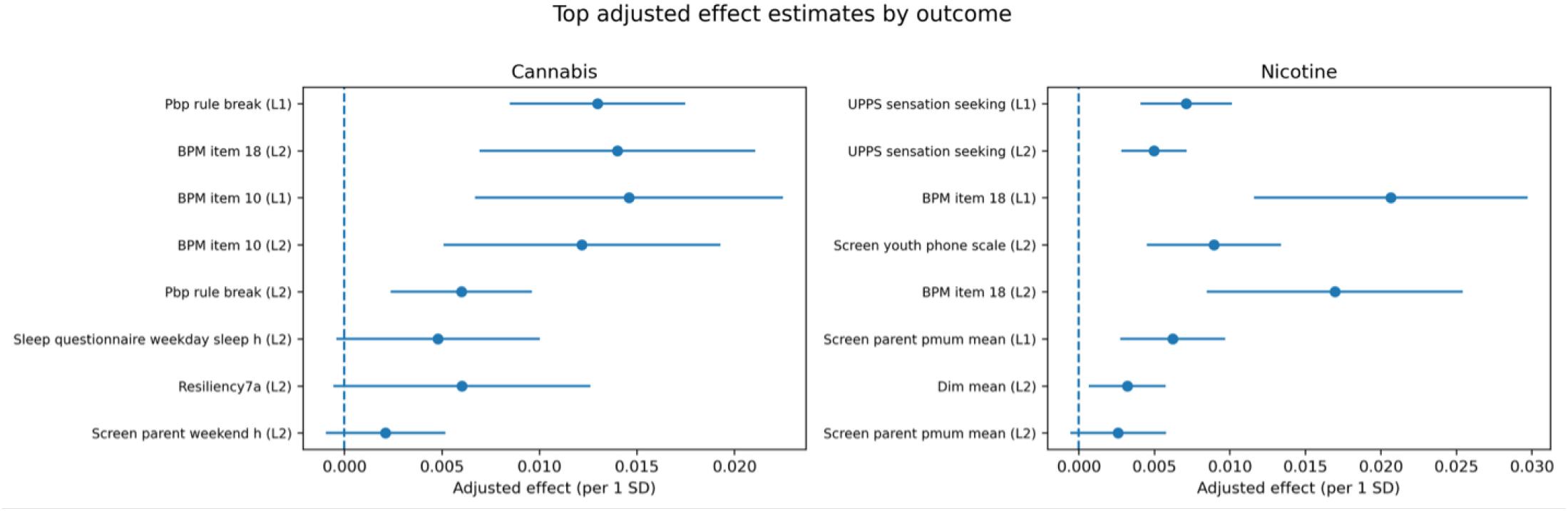
Top DML-style effect estimates by outcome. For each outcome, the figure shows the selected stable predictors with their adjusted DML-style effect estimates and 95% confidence intervals. Cannabis initiation was most clearly linked to rule-breaking behavior and behavioral items, whereas nicotine initiation was linked to sensation seeking, behavioral symptoms, and screen-related measures.

For alcohol initiation, the strongest stable predictor was youth resiliency, with a stability score of 0.580. Other alcohol-related predictors had lower stability, including another resiliency item and parent-reported rule-breaking behavior. This suggests that the stable graph for alcohol initiation was more selective and contained fewer strong incoming predictors than the cannabis and nicotine graphs.

For cannabis initiation, the strongest stable predictors were mainly behavioral and psychosocial variables. Parent-reported rule-breaking behavior showed very high stability, with scores of 0.915 and 0.880 across different lagged versions. Youth resiliency also showed high stability, with a score of 0.890. Other stable cannabis predictors included Brief Problem Monitor (BPM) behavioral items, weekday sleep duration, parent-reported weekend screen time, peer or social-context measures, and parent behavioral measures. This indicates that cannabis initiation was linked to a broader set of stable behavioral and environmental predictors.

For nicotine initiation, the strongest stable predictors included UPPS sensation seeking, screen-related parent and youth measures, BPM item 18, dimension mean scores, and Adult Self-Report (ASR) intrusive behavior. UPPS sensation seeking showed high stability at both lag 1 and lag 2, with stability scores of 0.705 and 0.690, respectively. Screen-related variables and BPM item 18 also showed moderate-to-high stability. These findings suggest that nicotine initiation was especially connected to impulsivity, behavioral symptoms, and screen- or family-environment variables.

Overall, the graph-discovery results showed both shared and outcome-specific patterns. Some predictors, such as rule-breaking behavior and resiliency, appeared across multiple outcomes, whereas others were more specific to cannabis or nicotine initiation.

### DML-Based Effect Estimation

#### Global Effect Estimates

After identifying stable graph edges, we estimated adjusted effects using a double machine learning (DML)-style residual-on-residual approach with cross-fitting. This step was used to estimate the direction and magnitude of association for selected stable predictors after adjustment for fixed covariates, including age, sex, site indicators, and genetic principal components.

The DML estimates should be interpreted as adjusted associations on an approximate risk-difference scale, not as definitive causal effects. Because the analysis was performed in a high-dimensional longitudinal setting, the confidence intervals are important for interpretation.

The strongest DML-style effects were observed for cannabis and nicotine initiation. Most effect sizes were modest in absolute magnitude, but several confidence intervals did not include zero.

For cannabis initiation, parent-reported rule-breaking behavior showed a positive association with later cannabis initiation. A 1-standard-deviation increase in rule-breaking behavior at lag 1 was associated with an estimated 0.013 increase in cannabis-initiation risk, with a 95% confidence interval from 0.008 to 0.017. BPM item 10 and BPM item 18 also showed positive associations with cannabis initiation. These findings suggest that behavioral problems were consistently linked to cannabis risk in both the stability-selection and DML analyses.

For nicotine initiation, BPM item 18 showed one of the largest positive effects. A 1-standard-deviation increase in BPM item 18 at lag 1 was associated with an estimated 0.021 increase in nicotine-initiation risk, with a 95% confidence interval from 0.012 to 0.030. UPPS sensation seeking was also positively associated with nicotine initiation at both lag 1 and lag 2. Screen-related variables, including the youth phone screen scale and parent-reported PMUM mean, also showed positive associations.

Some predictors had confidence intervals that crossed zero, including several cannabis-related sleep or screen variables and ASR intrusive behavior for nicotine initiation. These estimates should be interpreted with greater caution.

### Outcome-Specific Effects

The outcome-specific DML results further supported the idea that cannabis and nicotine initiation had different predictor profiles.

For cannabis initiation, the main positive predictors were parent-reported rule-breaking behavior and BPM behavioral items. Rule-breaking behavior was selected with high stability and also showed a positive adjusted effect. BPM item 10 and BPM item 18 also showed positive effects. These results suggest that cannabis initiation was most strongly linked to behavioral dysregulation and rule-breaking patterns in the current effect-estimation results.

For nicotine initiation, the strongest positive effects were observed for BPM item 18, UPPS sensation seeking, the youth phone screen scale, and the parent-reported PMUM mean. This suggests that nicotine initiation was related to impulsivity, behavioral symptoms, and screen- or parent-reported media-use patterns.

In the displayed DML top-effect tables, the strongest retained effects were mainly observed for cannabis and nicotine initiation. Therefore, the DML section focuses on these two outcomes unless additional alcohol or any-substance effect estimates are included in the final figures or supplementary tables.

### Summary of Key Findings

Together, the graph-discovery and DML analyses showed three main findings.

First, the bootstrap graph-discovery procedure identified several stable longitudinal relationships, especially for cannabis and nicotine initiation. Second, the most stable predictors were mainly behavioral and environmental variables, including rule-breaking behavior, sensation seeking, resiliency, screen-related measures, and family- or parent-related variables. Third, the DML-style estimates showed modest but positive adjusted effects for several of these predictors,especially rule-breaking behavior for cannabis initiation and sensation seeking, BPM item 18, and screen-related measures for nicotine initiation.

Overall, these results support the conclusion that substance-use initiation during adolescence is linked to a combination of shared and outcome-specific behavioral and environmental risk factors. In contrast, polygenic risk score variables were less consistently selected in the current graph-discovery results and did not appear as the dominant predictors in the displayed DML effect estimates.

## Discussion

### Main Findings

In this study, we used a longitudinal framework to examine genetic, behavioral, family, and environmental predictors of adolescent substance-use initiation. We combined several complementary approaches, including time-varying survival models, marginal structural model analyses, machine-learning prediction, graph discovery with stability selection, and DML-style effect estimation. Across these analyses, the most consistent signals came from behavioral and environmental predictors rather than polygenic risk scores.

The graph-discovery results showed that several predictors were repeatedly selected across bootstrap samples, suggesting that the learned relationships were not completely driven by random variation. The strongest stable edges were mainly related to behavioral symptoms, rule-breaking behavior, sensation seeking, resiliency, sleep, screen-related measures, and family- or parent-related factors. In contrast, polygenic risk score variables were less consistently selected and did not appear as dominant predictors in the current graph-discovery study.

The DML-style effect-estimation results further supported this pattern. Although the estimated effects were generally modest, several behavioral and environmental predictors showed positive associations with later substance-use initiation. For cannabis initiation, parent-reported rule-breaking behavior and BPM behavioral items showed the clearest positive associations. For nicotine initiation, BPM item 18, UPPS sensation seeking, the youth phone screen scale, and parent-reported media-use measures showed stronger effects. These results suggest that adolescent substance-use initiation is more closely related to behavioral and environmental risk patterns than to genetic liability alone in the current data.

### Shared and Substance-Specific Pathways

The results also suggest that substance-use initiation is influenced by both shared and substance-specific pathways. Some predictors appeared across multiple outcomes, suggesting that they may reflect broader vulnerability to substance use. These included rule-breaking behavior, resiliency-related measures, sleep-related variables, and family or environmental measures. These factors may capture general behavioral regulation, family context, and psychosocial environment, which can influence risk for more than one type of substance use.

At the same time, the strength and pattern of predictors differed by substance. Cannabis initiation showed strong and stable links with parent-reported rule-breaking behavior and other behavioral symptoms. Nicotine initiation was more strongly linked to sensation seeking, BPM behavioral items, and screen-related variables. Alcohol initiation showed fewer highly stable predictors in the current graph-discovery results, suggesting either a weaker detectable signal, a different risk structure, or less statistical power for the strongest edges after filtering.

These outcome-specific patterns are important because they suggest that adolescent substance use should not be treated as a single phenotype. Although alcohol, nicotine, cannabis, and any-substance initiation share some risk factors, each outcome may also have distinct predictors. This supports the use of multi-outcome and time-varying models because they can capture both common and substance-specific risk pathways.

### Interpretation of Effect Sizes

The DML-style effect estimates were modest in size. Most estimates were small on the risk-difference scale, even when the direction of association was consistent. This is expected in adolescent substance-use research, in which initiation is influenced by many small effects across individual, family, peer, school, and broader environmental domains.

Although the effect sizes were not large, they may still be meaningful. A small effect for one predictor may become important when it is part of a broader risk profile. For example, rule-breaking behavior, sensation seeking, sleep problems, and screen-related measures may each contribute modestly, but together they may help identify adolescents with a higher risk of later substance-use initiation.

However, these estimates should be interpreted carefully. The graph-discovery results identify stable longitudinal relationships, and the DML analysis estimates adjusted associations for selected predictors. These methods improve robustness compared with simple cross-sectional associations, but they do not prove definitive causality. Unmeasured confounding, measurement error, and changes in exposure over time may still affect the results. Therefore, the findings should be interpreted as evidence of reproducible longitudinal risk patterns rather than direct causal proof.

### Modifiable Risk Factors

A key finding of this study is that many of the strongest predictors are potentially modifiable. Behavioral symptoms, sensation seeking, sleep patterns, screen use, parental monitoring, and family environment are all areas that may be targeted through prevention or early intervention. This is important because polygenic risk score variables are not directly modifiable, whereas behavioral and environmental risk factors may be more useful for designing practical prevention strategies.

For example, the consistent association between rule-breaking behavior and cannabis initiation suggests that early behavioral dysregulation may be an important warning sign. Similarly, the association between sensation seeking and nicotine initiation suggests that prevention programs may need to address impulsivity, reward seeking, and risk-taking tendencies. Screen-related and parent-reported media-use variables may also reflect broader routines, supervision, or family-context factors that contribute to substance-use risk.

These findings do not mean that genetic risk is unimportant. Genetic liability may still contribute to adolescent substance use through complex pathways, including indirect effects through behavior, family environment, or peer selection. However, in the current analyses, polygenic risk score variables were weaker and less consistent than behavioral and environmental predictors. This suggests that, at least in this sample and with the available polygenic risk score measures, behavioral and environmental factors provided stronger signals for near-term substance-use initiation.

### Strengths

#### This study has several strengths

First, it used longitudinal data, allowing predictors and outcomes to be examined over time rather than only at a single time point. Second, it compared multiple analytic approaches, including survival analysis, marginal structural models, machine learning, graph discovery, and DML-style effect estimation. The consistency of findings across these different methods increases confidence in the overall pattern of results. Third, the study examined several substance-use outcomes rather than focusing on only one. This allowed us to identify both shared and substance-specific predictors. Fourth, the graph-discovery analysis used bootstrap stability selection, which reduced reliance on a single learned graph and helped prioritize more reproducible edges. Finally, the DML-style analysis provided adjusted effect estimates for selected stable predictors, helping to move from graph discovery toward more interpretable effect estimation.

### Limitations

This study also has several limitations. First, although the analysis used longitudinal data and causal-discovery tools, the findings should not be interpreted as definitive causal effects. Residual confounding and unmeasured variables may still influence the results. Second, some substance-use outcomes may have fewer events or weaker signals, which can reduce statistical power and make stable-edge detection more difficult.

Third, many predictors were self-reported or parent-reported. These measures may contain reporting bias or measurement error. Fourth, the polygenic risk score variables may not fully capture genetic liability for substance-use initiation, especially if the discovery genome-wide association studies did not match the developmental stage, ancestry composition, or phenotype definition of the current sample. Fifth, the current results were based on the available ABCD follow-up period, and longer follow-up may reveal stronger or different patterns as more participants initiate substance use.

Finally, the graph-discovery and DML analyses required filtering and selection steps. These steps improve interpretability, but they may also exclude some weaker or indirect pathways. Therefore, the results should be viewed as a prioritized set of robust associations rather than a complete map of all possible mechanisms.

### Conclusion

In conclusion, this study identified reproducible longitudinal predictors of adolescent substance-use initiation using a multi-step framework that combined graph discovery, stability selection, and DML-style effect estimation. Across analyses, behavioral and environmental predictors were more consistent than polygenic risk scores. Rule-breaking behavior, sensation seeking, behavioral symptoms, screen-related measures, sleep, resiliency, and family- or parent-related factors showed the strongest and most interpretable signals.

The findings suggest that adolescent substance-use initiation is shaped by both shared and substance-specific pathways. Cannabis initiation was most clearly linked to rule-breaking behavior and behavioral symptoms, whereas nicotine initiation was linked to sensation seeking, behavioral symptoms, and screen-related measures. Polygenic risk score variables showed weaker and less consistent associations in the current analyses.

Overall, these results highlight the importance of modifiable behavioral and environmental risk factors in adolescent substance-use initiation. They also support the value of longitudinal, multi-method approaches for identifying early risk patterns. Future studies with longer follow-up, more substance-use events, and external validation will be important for confirming these findings and evaluating whether the identified predictors can improve prevention and early-intervention strategies.

## Data Availability

This study uses data from the Adolescent Brain Cognitive Development (ABCD) Study (abcdstudy.org), held in the NIMH Data Archive (NDA). The ABCD data version used was 5.1. The study is supported by the National Institutes of Health (NIH) and additional federal partners under multiple award numbers, including U01DA041048 and U01DA050987. The full list of funders is available at abcdstudy.org/federal-partners.html.

## Data availability

### Code

The analysis code and scripts used in this study are publicly available at: https://github.com/mw742/ABCD-CausalML

### Data

This study uses data from the Adolescent Brain Cognitive Development (ABCD) Study (https://abcdstudy.org), available through the NIMH Data Archive (NDA). The ABCD data release used was version 5.1. The study is supported by the National Institutes of Health (NIH) and additional federal partners under multiple award numbers, including U01DA041048 and U01DA050987. A full list of funders is available at https://abcdstudy.org/federal-partners.html.

## Author contributions statement

Mengman Wei conceived the study, designed the analytical framework, performed data processing, statistical analyses, and computational modeling, and drafted the manuscript. Mengman Wei also carried out code implementation, data curation, and interpretation of the results. Lasya Yadlapati contributed to data curation, visualization, and manuscript discussion. Qian Peng provided supervision, overall guidance, resource support, and funding acquisition. All authors contributed to the discussion and revision of the manuscript.

## Preprint Notice

This manuscript is a preprint and has not yet undergone peer review. The content is shared to disseminate findings and establish a precedent. Additional analyses and revisions may be incorporated in future versions.

## References

1. Ronald C Kessler, G Paul Amminger, Sergio Aguilar-Gaxiola, Jordi Alonso, Sing Lee, and T Bedirhan Üstün. Age of onset of mental disorders: a review of recent literature. Current opinion in psychiatry, 20(4):359–364, 2007.

2. ReJoyce Green, Bethany J Wolf, Andrew Chen, Anna E Kirkland, Pamela L Ferguson, Brittney D Browning, Brittany E Bryant, Rachel L Tomko, Kevin M Gray, Louise Mewton, et al. Predictors of substance use initiation by early adolescence. American journal of psychiatry, 181(5):423–433, 2024.

3. Iliyan Ivanov, Muhammad A Parvaz, Eva Velthorst, Riaz B Shaik, Sven Sandin, Gabriela Gan, Philip Spechler, Matthew D Albaugh, Bader Chaarani, Scott Mackey, et al. Substance use initiation, particularly alcohol, in drug-naive adolescents: possible predictors and consequences from a large cohort naturalistic study. Journal of the American Academy of Child & Adolescent Psychiatry, 60(5):623–636, 2021.

4. Terry L Jernigan, Sandra A Brown, and Gayathri J Dowling. The adolescent brain cognitive development study. Journal of research on adolescence: the official journal of the Society for Research on Adolescence, 28(1):154, 2018.

5. National Institutes of Health (NIH). Adolescent Brain Cognitive Development (ABCD) Study. https://abcdstudy.org, n.d. Accessed: 2026-04-15.

6. Norbert Wiener. The theory of prediction. modern mathematics for engineers. New York, 165(6), 1956.

7. Clive WJ Granger. Investigating causal relations by econometric models and cross-spectral methods. Econometrica: journal of the Econometric Society, pages 424–438, 1969.

8. Robert Tibshirani. Regression shrinkage and selection via the lasso. Journal of the Royal Statistical Society Series B: Statistical Methodology, 58(1):267–288, 1996.

9. Arthur E Hoerl and Robert W Kennard. Ridge regression: Biased estimation for nonorthogonal problems. Technometrics, 12(1):55–67, 1970.

10. Trevor Hastie. The elements of statistical learning: data mining, inference, and prediction, 2009.

11. Nicolai Meinshausen and Peter Bühlmann. Stability selection. Journal of the Royal Statistical Society: Series B (Statistical Methodology), 72(4):417–473, 2010.

12. Hui Zou and Trevor Hastie. Regularization and variable selection via the elastic net. Journal of the Royal Statistical Society Series B: Statistical Methodology, 67(2):301–320, 2005.

13. Alexandre Belloni, Victor Chernozhukov, and Christian Hansen. Inference on treatment effects after selection among high-dimensional controls. Review of Economic Studies, 81(2):608–650, 2014.

14. Mark J Van Der Laan and Daniel Rubin. Targeted maximum likelihood learning. 2006.

15. Sören R Künzel, Jasjeet S Sekhon, Peter J Bickel, and Bin Yu. Metalearners for estimating heterogeneous treatment effects using machine learning. Proceedings of the national academy of sciences, 116(10):4156–4165, 2019.

16. Philipp Bach, Victor Chernozhukov, Malte S Kurz, and Martin Spindler. Doubleml-an object-oriented implementation of double machine learning in python. Journal of Machine Learning Research, 23(53):1–6, 2022.

17. Mengman Wei and Qian Peng. Time-varying environmental and polygenic predictors of substance use initiation in youth: A survival and causal modeling study in the abcd cohort. arXiv preprint arXiv:2604.07368, 2026.

18. Jeffrey M Wooldridge. Econometric analysis of cross section and panel data. MIT press, 2010.

19. Judith D Singer and John B Willett. Applied longitudinal data analysis: Modeling change and event occurrence. Oxford university press, 2003.

20. Victor Chernozhukov, Denis Chetverikov, Mert Demirer, Esther Duflo, Christian Hansen, Whitney Newey, and James Robins. Double/debiased machine learning for treatment and structural parameters, 2018.

21. Leo Breiman. Random forests. Machine learning, 45(1):5–32, 2001.

22. Kung-Yee Liang and Scott L Zeger. Longitudinal data analysis using generalized linear models. biometrika, pages 13–22, 1986.

